# A Compendium of manually annotated genetic variants for Alkaptonuria-AKUHub

**DOI:** 10.1101/2023.02.21.23286262

**Authors:** S Akila, T.C Arun Kumar, S Shofia, S Srividya, N Suriyamoorthy, R Vijhayalakshmi, Vinod scaria, Ratnesh Bhai Mehta

## Abstract

Alkaptonuria or ‘black urine disease’ is a rare autosomal recessive disorder caused by dysfunctional homogentisate 1,2-dioxygenase (HGD) gene (3q13.33) leading to accumulation of homogentisic acid in the body. This inborn error in metabolism of phenylalanine and tyrosine causes accumulation of homogentisic acid leading to ochronosis, pigmentation in the sclera, ear cartilage, mitral valve calcification and osteoarthropathy. Advances in sequencing technologies have helped us to map genetic variants associated with alkaptonuria in diverse populations and regions. Currently, no centralized resource of all the reported actionable variants with uniformity in annotation exists for the HGD gene. We have compiled HGD exonic variants from various data sources and systematically annotated their pathogenicity according to American College of Medical Genetics and the Association of Molecular Pathologists (ACMG/AMP) variant classification framework. A total of 1686 exonic variants were catalogued and manually curated, creating one of the most comprehensive Alkaptonuria variant databases (AKUHub) which is publicly available.

## 2. Introduction

Alkaptonuria is a rare autosomal recessive genetic disorder caused by mutation in HGD gene, discovered by Sir Archibald Edward Garrod in 1902. Garrod had defined alkaptonuria as one of the inborn errors of metabolism in his Croonia lectures^1^. The primary cause of homogentisate 1,2-dioxygenase (HGD) deficiency is mutations in the HGD gene and is mapped to chromosome 3q13.33. HGD is involved in the breakdown of the amino acids phenylalanine and tyrosine, which are essential components of many proteins in the body. In people with HGD deficiency, the enzyme is either partially or completely non-functional, leading to the accumulation of a toxic intermediate called homogentisic acid (HGA) in the body^2^. This buildup of homogentisic acid can cause a range of symptoms including joint pain, arthritis and discoloration of the skin and eyes, ochronotic osteoarthropathy^3^ and ochronosis^4^ (bluish black or grayish blue pigmentation in connective tissue). Ochronosis mostly occurs after age of 30 years and this disorder may go unnoticed till adulthood because other symptoms usually do not appear until the age of 30. Other manifestations can include pigmentation in the sclera, ear cartilage and skin of the hands, aortic or mitral valve calcification or regurgitation, renal stones, prostate stones, and hypothyroidism. The biochemical diagnosis of alkaptonuria is based on the detection of substantial amounts of HGA (usually 1-8 g per day) in the urine^3^. Worldwide, the prevalence of Alkaptonuria is 1 in 250,000-100,000 live births^5^. The high prevalence rate has been reported in the regions of Slovakia and Dominican Republic, affecting 1 in 19,000 people^6^. Several cases have been reported in India but prevalence rate in the population remains unclear as of now. Specific communities for example, Gypsy population of Narikuravas residing around Vellore districts of Tamil Nadu, India have been reported to have high prevalence and significant founder mutation effect^7^ in the HGD gene.

Even though Alkaptonuria is not life-threatening, it affects the quality of life. Disease severity varies between affected individuals, even between siblings, and increases with age due to the accumulation of HGA. Currently there is no specific treatment or cure. Symptomatic treatment and lifestyle modifications can help to manage the symptoms without compromising the quality of life. In addition to the development of a potentially disease-modifying therapy, clinical evaluation of the condition has been improved. Early diagnosis helps in timely spinal surgery and arthroplasty which are the major treatment approaches at present and restricting dietary protein which can lower the levels of tyrosine and phenylalanine in the body, thereby slowing down the development of disease. Periodic surveillance to detect and treat the complications at the earliest is important. Alkaptonuria follows Mendelian autosomal recessive pattern, wherein if both parents are known to be heterozygous (carrier) for an HGD pathogenic variant, each sibling of an affected individual has 25% chance of being affected, a 50% chance of being an asymptomatic carrier, and a 25% chance of being unaffected and this implies the importance of carrier testing for at-risk relatives.

Single gene testing can establish the carrier status in proband along with the additional recurrence risk assessment for family members. Once the HGD pathogenic variants have been identified in an affected family member, prenatal testing and preimplantation genetic testing/ genetic counseling is required to provide reliable recurrence risk assessment^3^.

Studies focusing on identifying pathogenic variants have shown remarkable allelic heterogeneity despite low prevalence^8^. Some of the mutations are more prevalent in certain populations or ethnicities while some of them have been reported worldwide^9^. Understanding the underlying pathogenicity of the variant is critical for the prevention of recurrence in offspring and for the elucidation of disease mechanisms. However, the lack of a uniform mechanism to identify the pathogenicity of genetic variations for clinical interpretation prevents widespread use of such knowledge in clinical settings. ACMG/AMP released guidelines on the annotation of genetic sequence variants that provide a uniform framework for systematic integration of evidence on each variant and classifies them based on the evidence obtained to infer their pathogenicity^10^. Systematic curation and annotation of HGD genetic variations can meet the need for a therapeutically relevant resource for alkaptonuria diagnosis. In the current manuscript we have systematically curated and cataloged all the protein coding HGD gene variants in accordance with ACMG/AMP framework to create the most comprehensive curated online database for alkaptonuria. A total of 1686 exonic variants from various literatures and databases for HGD gene were manually collected and annotated. This database is publicly accessible on https://www.zifogenomics.com/AKUHub and serves as a central point for clinicians and geneticists working on alkaptonuria.

## 3. Materials and Methods

### 3.1 Variant Consolidation

HGD gene variant database was created by systematically compiling the variants from various public variant databases namely ClinVar^11^, Leiden Open Variation Database (LOVD)^12^, Mastermind^13^, and Institute of Genomics & Integrative Biology (IGIB) South Asian Genomes & Exome (SAGE) database^14^, gnomAD^15^, dbSNP^16^, IndiGEN^17^ respectively. Additionally, variants were also compiled by manually carrying out literature search in search engines like Google Scholar, PubMed etc. using various combination of keywords like “HGD and Variants”, “Alkaptonuria and Variants”. Information pertaining to HGD variants like HGVSc, HGVSp and rsID prior to December 2022 were extracted from various public data sources (Variant Validator^18^ & VEP^19^) and was systematically captured in Microsoft excel [Version 2201 Build 16.0.14827.20180].

Variant Validator (Version 2.2.0)^18^ was used to align the Gene, Chromosomal location (coding position), Ref and Alt alleles and amino acid of the variant as per Human Genome Variation Society (HGVS)^20^ guidelines in addition to carry out syntax checking and validation. All information pertaining to variants were in the lines of Genome Reference Consortium Human build 38 (GRCh38). Variants were consolidated into a unique list for variant annotation purpose.

### 3.2 Variant Annotation

Variants were re-annotated as pathogenic, likely pathogenic, benign, likely benign and variant of uncertain significance (VUS) according to the joint consensus recommendation of the ACMG/AMP guidelines as described ^21^. In brief, variant details like gene name, chromosomal location / region, were collected from variant validator and rsID, allele frequency and variant type were collected from variant effect predictor (VEP)^19^. VEP reported NM_000187.4 refseq transcript for HGD gene. Exonic variants were filtered out for further pathogenicity classification and for the creation of HGD database. Variants were manually re-annotated based on information collected from computational predictive programs (Mutation Taster^22^-Polyphen^23^-CADD^24^), population dataset, disease-specific and sequence databases and literature in following order. (Figure 1)

**Figure 1:**
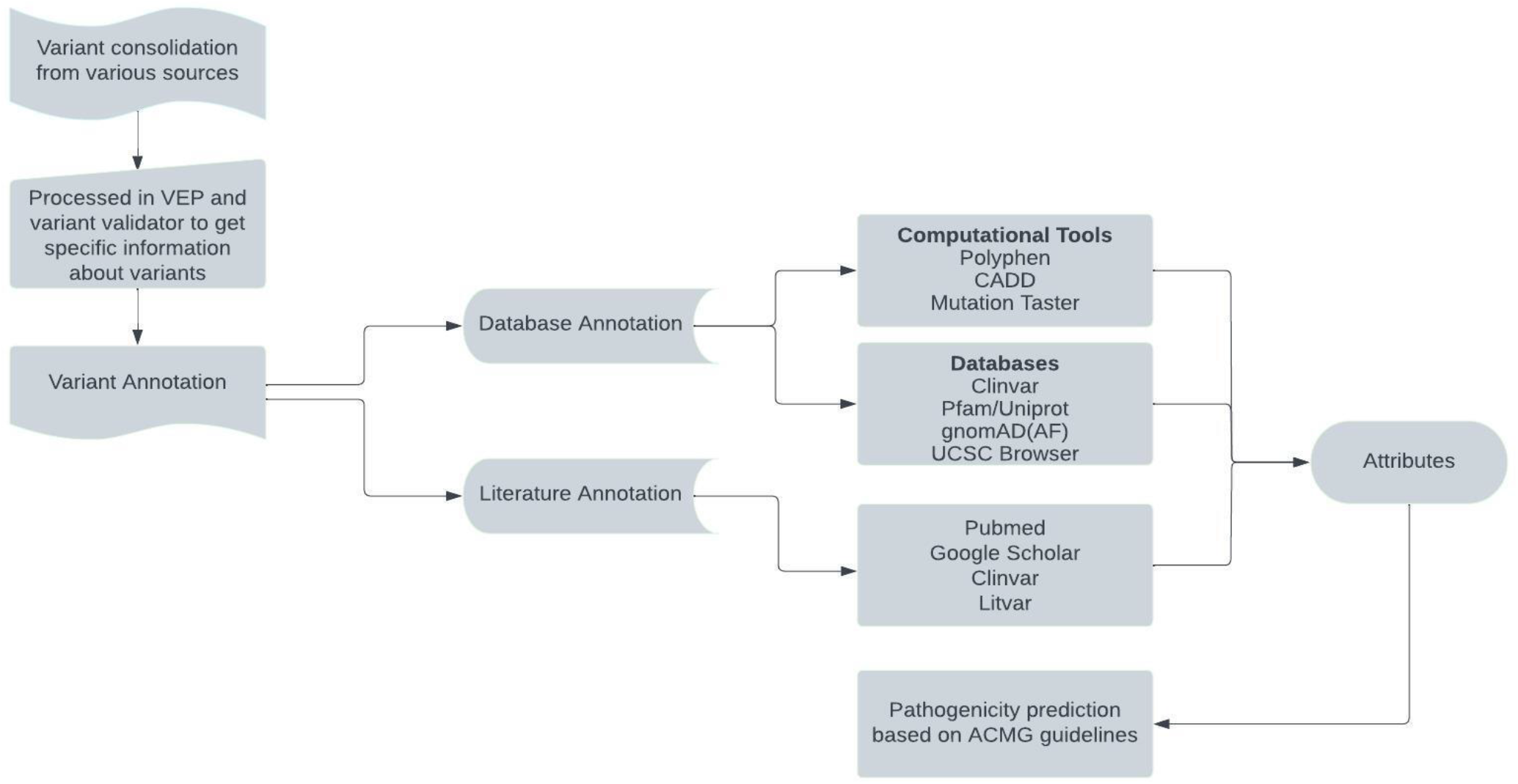
Process flow implemented for re-annotating variants as per the ACMG guidelines.

### Computational (*In Silico*) Predictive Tools

Variants were attributed either PP3 or BP4 if at least two of the *in-silico* tools namely Mutation Taster, Polyphen 2.0, CADD supported the deleterious effect or having no impact on the protein function respectively.

### Database annotation

Publicly available databases, predictive softwares, and published data obtained from relevant papers were used to provide attribute/criteria specifications. Missense variants were classified as PM1, if the variant was found in mutational hotspot/function domain as given in Pfam database^25^. BP1 is assigned for missense variant in a gene in which truncating variants are primarily known to cause the disease. PP2 is assigned if the missense variant in a gene that has a low rate of benign missense variation and where missense variants are a common mechanism of disease. The allele frequency of reported variants was explored in gnomAD / 1000 genome projects and attributes were assigned as BA1 (>0.05), BS1 (0.01 – 0.05), PM2 (<0.0005), BS2(observed in healthy adults) based on the reported Allele frequency^15^. PP5 was assigned if reputable source such as ClinVar reported the variant as pathogenic whereas BP6 is assigned for the benign evidence^11^. BP7 was assigned for all synonymous variants and PVS1 was assigned for frameshift, stop lost, start lost and stop gain variants. For indels, BP3 and PM4 were attributed based on the presence or absence of repeat region from UCSC browser^26^ respectively.

#### 3.2.1 Literature Annotation

The relevant articles associated with variants were retrieved from web sources like - Google scholar, PubMed, Litvar^27^, ClinVar using specific search combination such as “Gene name and Nucleotide position”, “Gene name and Amino acid change”, “Gene name and rsID”. Articles specific to the variant were glanced over and every article was manually annotated to assign relevant ACMG literature attribute information. The evidence supporting the attributes were recorded along with the Unique identifier (DOI, PMID) of the article. The attributes were assigned to each variant as described (Table 1).

**Table 1:** Literature attributes and their description

| Parameters | ACMG Attributes | Description |
| --- | --- | --- |
| Functional Study | BS3 | Well established *in vitro/in vivo* functional studies which have no effect on the protein |
|  | PS3 | Well established *in vitro/in vivo* functional studies supportive of a deleterious or disease-causing effect |
| Odds ratio | PS4 | Prevalence in cases over controls (odd's ratio >5) |
| Family segregation | BS4 | Lack of segregation with the disease in affected family members |
|  | PP1 | Co segregation with disease in multiple affected family members |
|  | PP4 | Patient's phenotype or family history is highly specific for a disease with a single genetic etiology |
| Cis/Trans variants | BP2 | Observed in *Cis/Trans* with a Pathogenic variant |
|  | PM3 | Observed in *Trans* with a pathogenic variant (Recessive disorder) |
| Alternate causation of disease | BP5 | Given in case of an alternate causation of disease |
| De novo Mutation | PM6 | De novo without paternity or maternity confirmed |
|  | PS2 | De novo with paternity or maternity confirmed |

### 3.3 Pathogenicity Prediction

All the variant specific attributes were compiled, and the pathogenicity was predicted using Genetic Variant Interpretation Online Tool developed by the University of Maryland^28^.

### 3.4 Database creation

AKUHub has been established in 2021 as a publicly accessible database, which aims to streamline the evaluation of HGD gene variants. It was developed with the intention of helping the clinicians and geneticists to assist ongoing clinical and molecular research in the field of AKU. This database lists around 1686 exonic variants updated as of December 2022. This web-based system was formatted and ported onto MySQL 5.7.33. HTML5, CSS3, Bootstrap was used to code this web interface. The backend was constructed using PHP 7.4.10, MySQL 5.7.33 and the server of this database was hosted using Apache2 server. Keep track of data processing, MySQL was used. jQuery and PHP 7.4.10 scripts were used with search query for retrieving the database information.

## 4. Results

Homogentisate 1,2-dioxygenase (HGD) is a single-copy gene that spans from 120,628,172 to 120,682,239 in the reverse strand of the genomic sequence on the long arm of chromosome 3 (3q13.33) (54,068 bp). The homogentisate oxidase enzyme, which is mostly involved in the liver and kidney mechanisms, is encoded by this gene, which has 14 exons with 445 amino acids in it. According to ACMG/AMP criteria, we have compiled variants from publicly available databases as well as from literatures to create the most comprehensive HGD exonic genetic variants database.

### 4.1 Variant consolidation

We collected 5460 variants from various data sources-ClinVar, Leiden Open Variation Database (LOVD), GenomeAsia29, GNomAD, Mastermind, Indigen, dbSNP and literature (Supplement table 1). The variants complied were found in exonic (2447), intronic regions (2841), untranslated regions (75) and splice sites (97) regions of the gene as per VEP (Figure 2). After removing duplicates, 1686 were unique exonic variants and were summarised based on variant type as Missense (1240), Synonymous (316), Frameshift (66), non-frameshift (64). We discovered 2 benign (0.12%), 262 Likely benign (15.54%), 170 Likely pathogenic (10.08%), 131 Pathogenic (8.42%) and 1121 variant of unknown significance (VUS) (66.48%) variants when we thoroughly re-annotated these 1686 variants as per the ACMG/AMP guidelines (Figure 3). In Figure 4, it can be observed that variants were spread across all 14 exons with exon 13 having the highest number of variants (n=228) accounting for 13.5% of all exonic variants.

**Figure 2:**
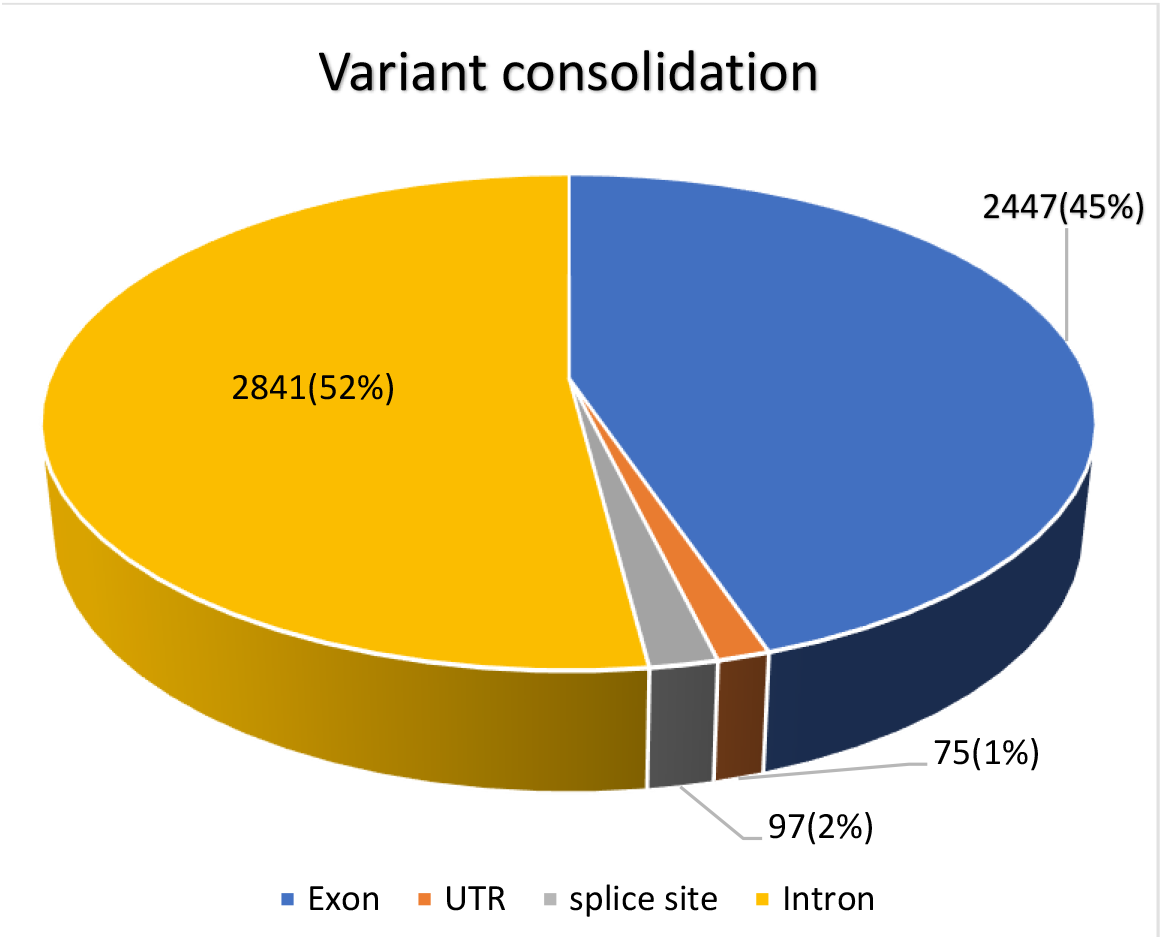
HGD Variant consolidation-variants were consolidated from various databases like ClinVar, Leiden Open Variation Database (LOVD), GenomeAsia, GNomAD, Mastermind, Indigen, dbSNP and literature.

**Figure 3:**
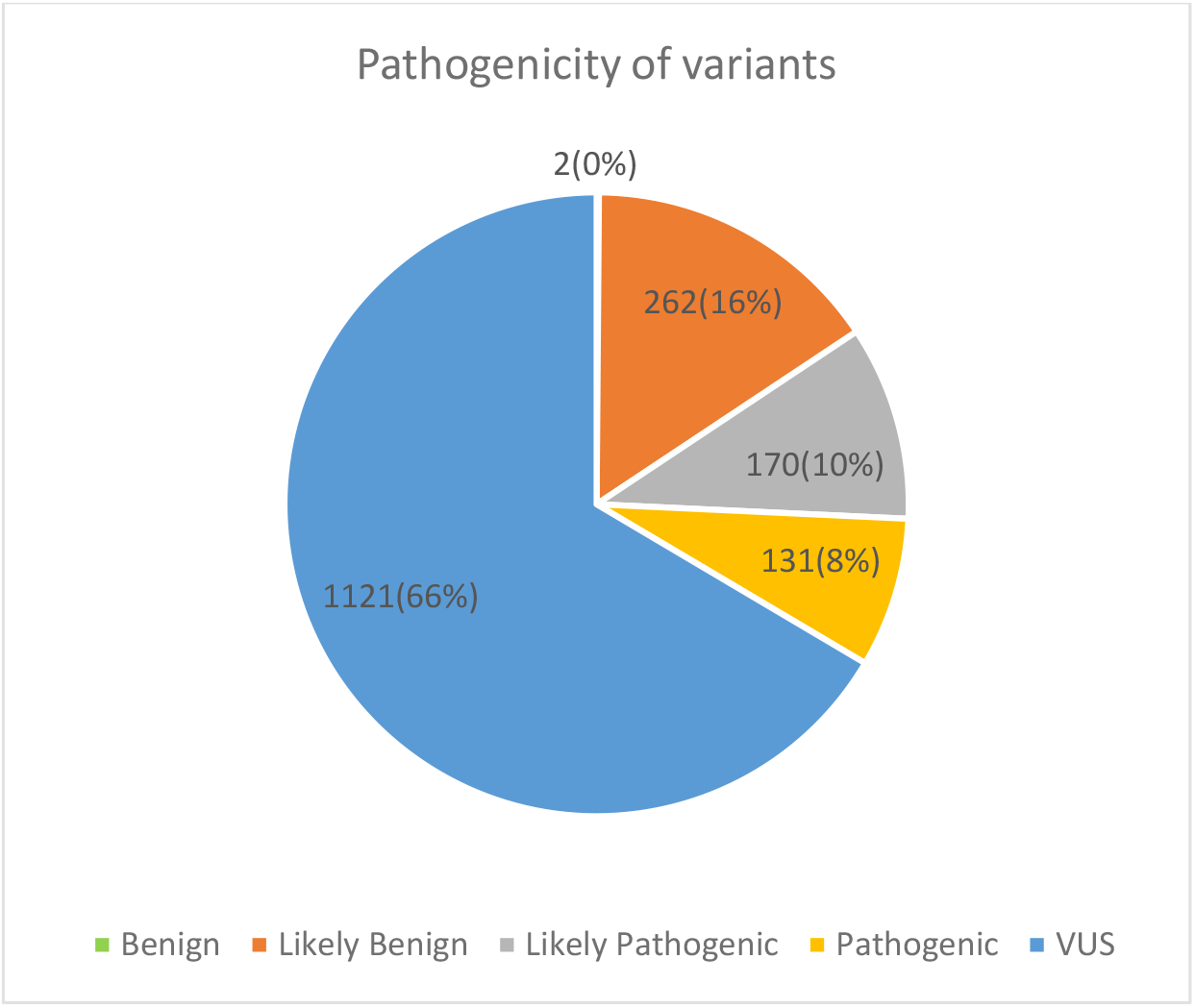
Pathogenicity of HGD exonic variants-When thoroughly re-annotating these 1686 variants using the conventional parameters, it was found that benign (0.12%), Likely benign (15.54%), Likely pathogenic (11.27%), Pathogenic (8.42%), and variant of uncertain significance (VUS) (64.65%) were all present.

**Figure 4:**
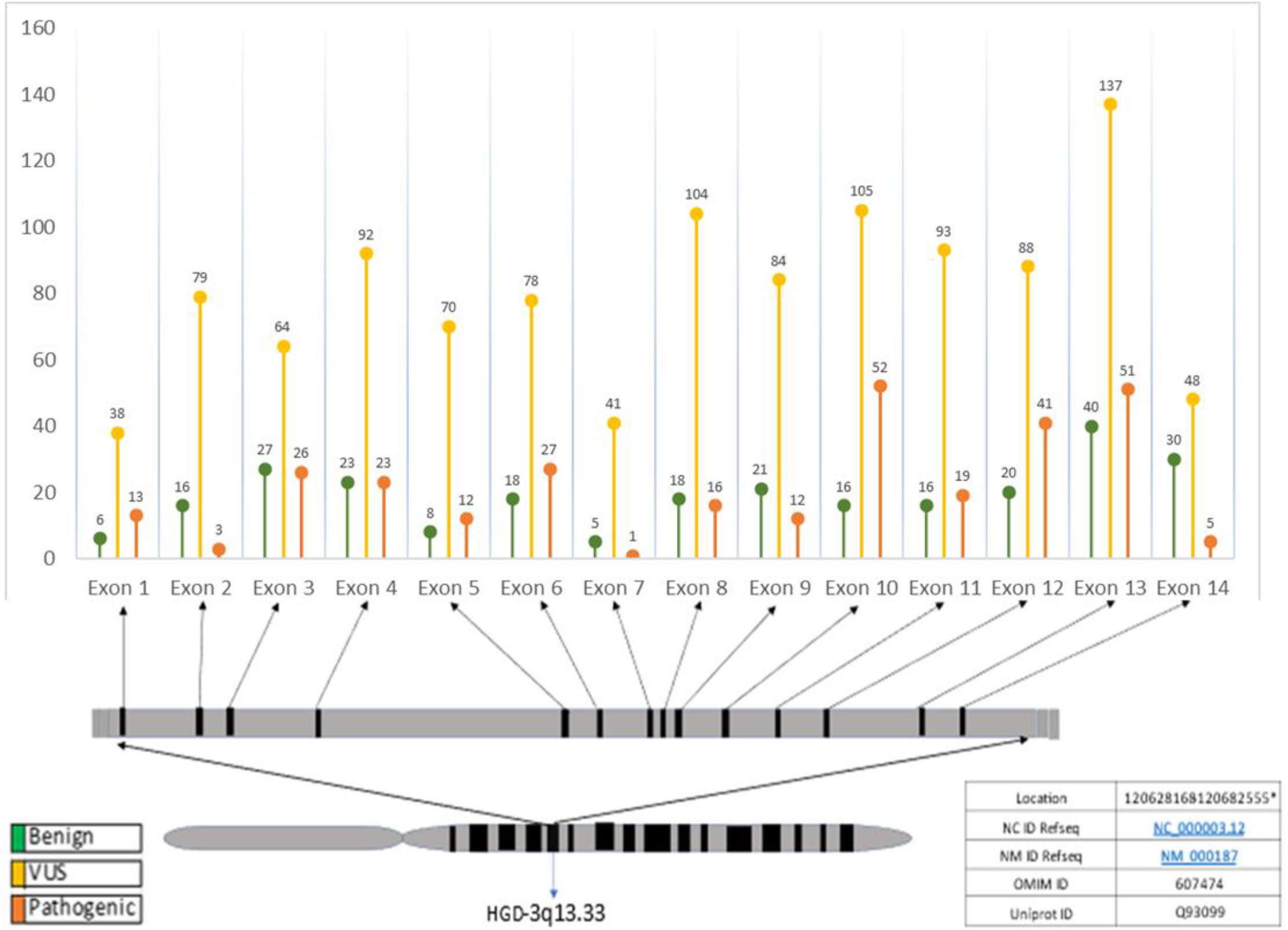
Variants distribution - Schematic representation of Variant distribution among the 14 exons. The majority of variations are found in exons 13 and 10.

### 4.2 Distribution of HGD variants based on geography

HGD exonic variants showed heterogenetic in respect to its frequency and distribution in different populations. Out of 1686 exonic variants, we were able to map the geographical distribution of 264 variants from scientific literatures. The variants were found to be distributed across 37 countries worldwide (Figure 5). Most of the developed countries like USA, UK, France, Spain reported highest number of HGD variants, however overall prevalence of these variants might be unexplored in the developing countries due to limited variant information. Pathogenic / likely pathogenic variants were primarily distributed across 33 countries.

**Figure 5:**
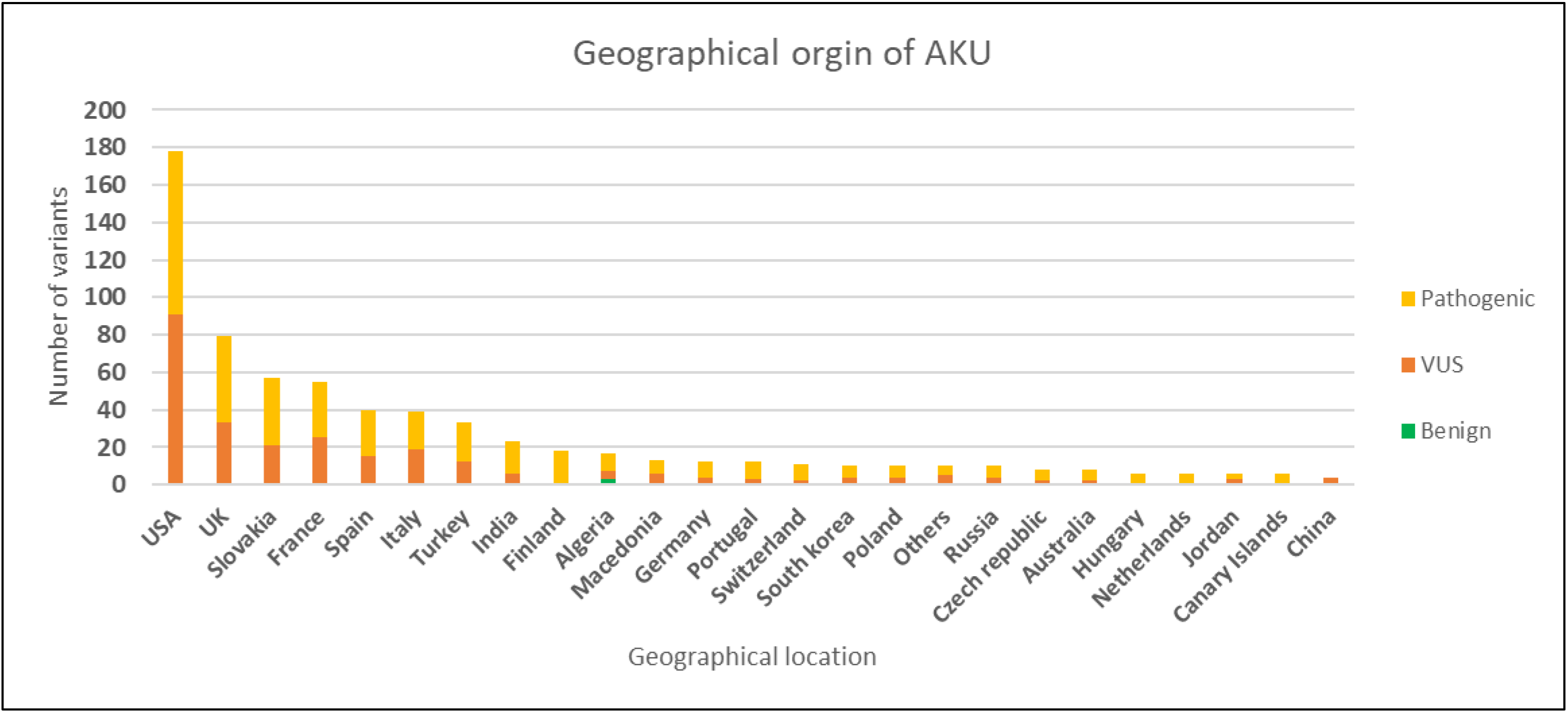
Geographical distribution of 264 pathogenic AKU variants previously reported in 37 countries worldwide.

The mutations c.1102A>G, c.481G>A, c.175del, and c.899T>G have been reported in over ten geographical locations. Among the mutations with known geographical distribution, we found that approximately 5% (12 out of 264) were exclusively present in the Indian population. These include c.362G>A, c.1019G>T, c.469G>T, c.504G>T, c.347_348delinsCC, c.347_348delinsCT and c.347_348inv which were classified as variants of uncertain significance (VUS). Meanwhile, c.504G>C, c.1039C>T, c.365_366delinsTA, c.365_366delinsTC and c.365_366delinsTG were pathogenic. Interestingly, despite these 12 variants being reported to be present in india from the scientific literature, our study revealed that none of these variants were included in Indigen database (as of 2020).

### 4.3 Impact of Pathogenicity on variant type and mutation class

We examined the types of mutation both at the nucleotide and protein level. The most frequent mutation class was observed in substitution (54.9%), followed by delins (38.8%), deletion (2.9%), inversion (2.1%), insertion (0.8%) and duplication (0.6%). Since the similar type of nucleotide change at the DNA level might result in various protein change, the variant types at the amino acid level were also examined. Missense was observed to be the most frequent variant type, representing about 73.5% of the exonic variants. The proportions of other variant types include synonymous (18.74%), frameshift (3.85%), missense (73.5%), stop gain (2.5%), stop loss (0.05%) and start lost (0.83%). Likely benign/Benign variants were present majorly in Synonymous. (Figure 6).

**Figure 6:**
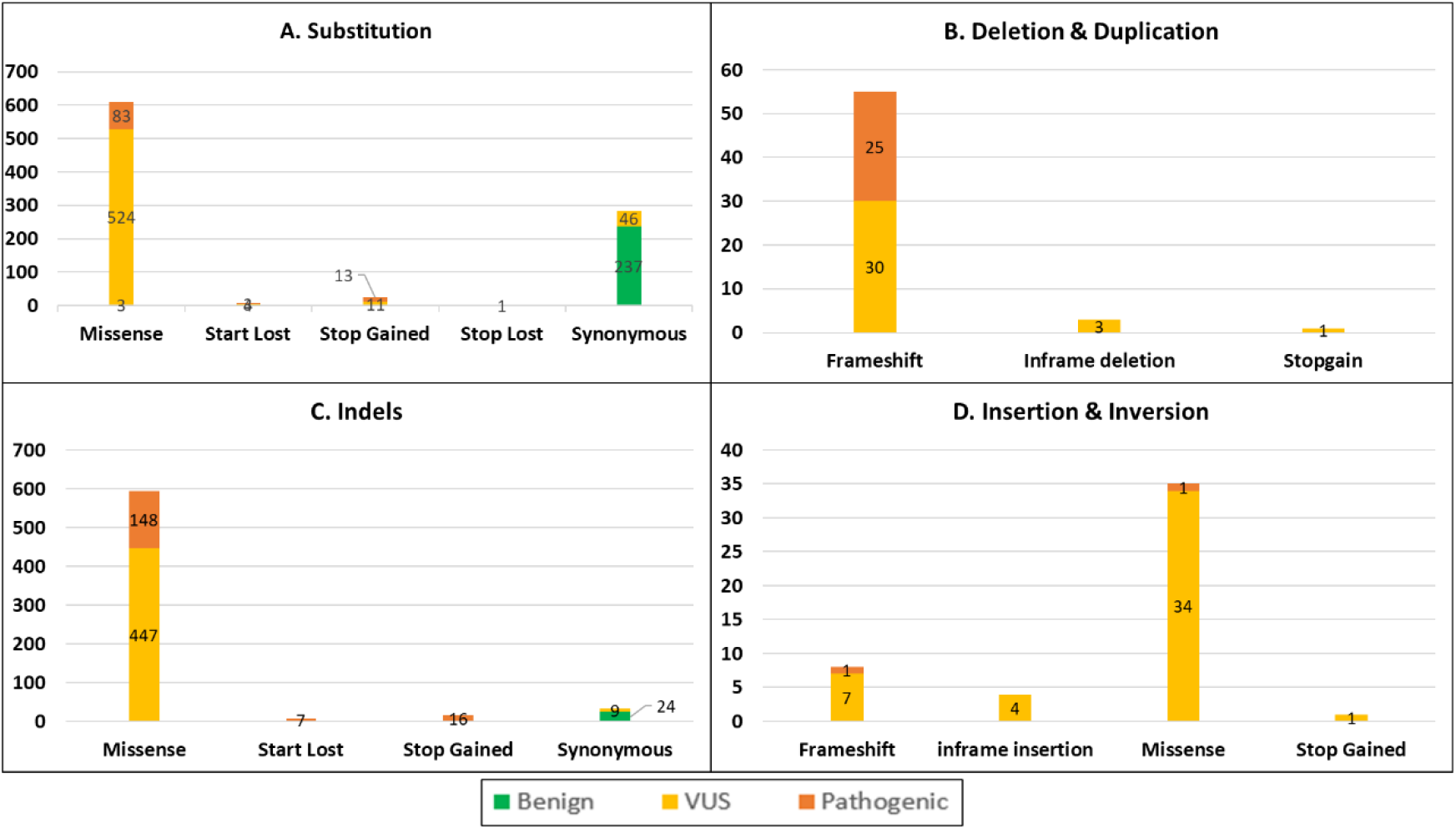
Impact of Pathogenicity on variant type and mutation class. The mutation class - A) Substitution B) Duplication & Deletion C) Indels D) Insertion & Inversion have been compared with variant type and pathogenicity.

The HGD protein is composed of two essential domains, HgmA_C and HgmA_N, that play a critical role in its enzymatic activity. Mutations in the HgmA_N domain of the HGD protein can impact protein folding and stability, leading to the accumulation of misfolded proteins that interfere with normal cellular processes. In the case of the HgmA_C, mutations can lead to a loss of enzymatic activity and the buildup of homogentisic acid in the body30.Our study found that the majority of variants identified fell within the HgmA_N domain (amino acid position: 5-273, exon 1-exon 11), with 68% of the 307 pathogenic variants located in this region (Figure 7).

**Figure 7:**
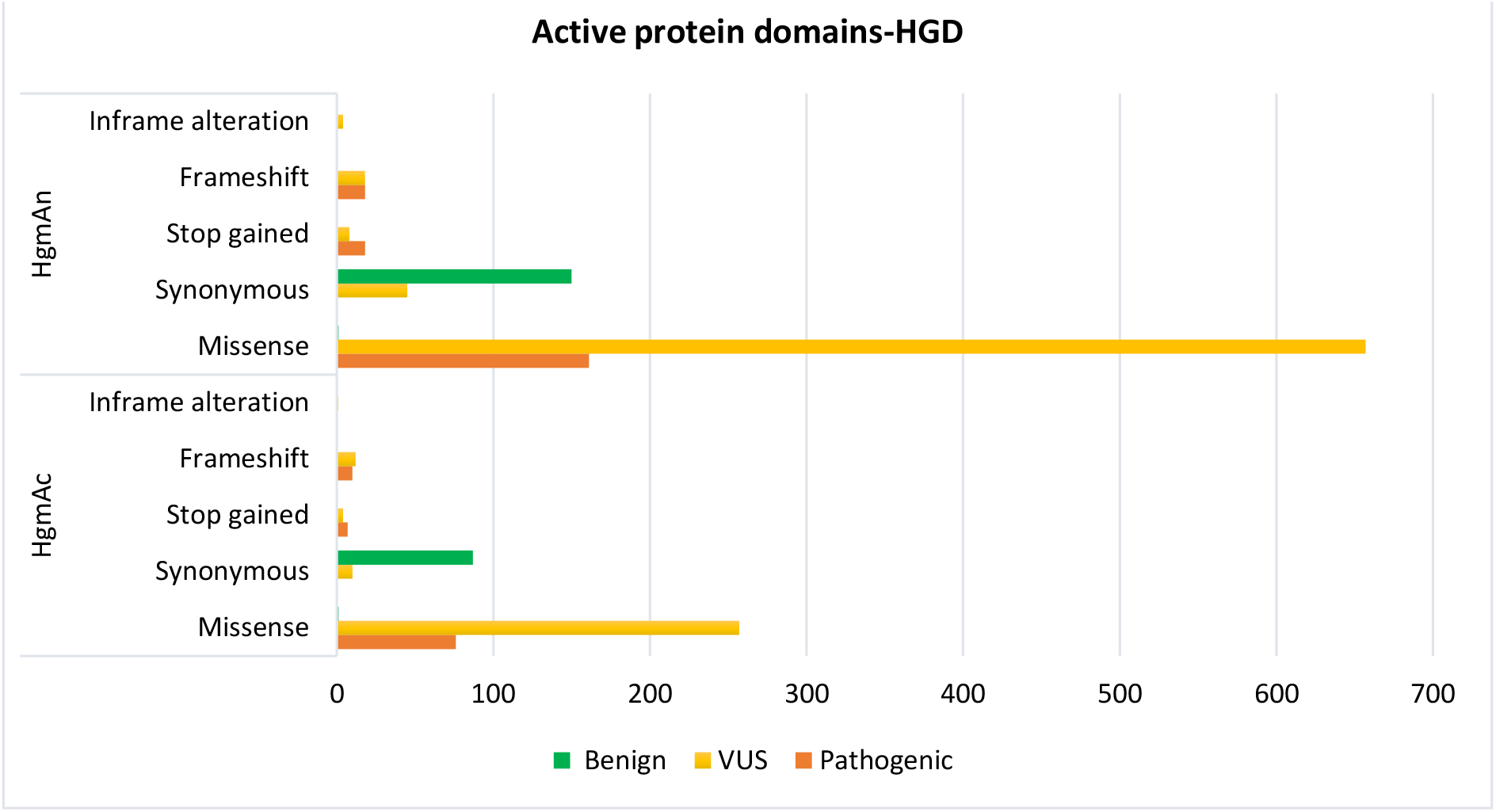
Pfam domain for HGD gene-The HGD’s structural complexes as seen via Pfam. Variant occurrences in the domain were represented together with their pathogenicity and variant type. Uniport reference number (HGD_HUMAN-Q93099)

### 4.4 ACMG attributes and its features

#### Functional assay

Variants of HGD gene are not fully explored in context to its functional activity. The scientific literature contained few functional assays limited to cell lines and yeast models, which we considered to be one of the key defining criteria for assigning the pathogenicity of variants. We found 11 pathogenic variants for which a well-established functional assay has been performed categorizing it as pathogenic (Supplement table 2).

#### *In silico* prediction

Using the computational tools-CADD algorithm, PolyPhen2 and mutation taster, we assigned in silico score for all the variants (that had enough evidence). Comparing with in silico scores, we found only four non-truncating missense variants (p.Asn278Asp, p.Arg307His, p.Pro308Arg, and p.Asp326Asn) were conflicting with the final pathogenicity determined using the ACMG standards (Supplement table 3).

#### Population frequency

Public sources like gnomAD and VEP had information on the population frequency for about 562 variants. Figure 8 highlights the prevalence of genetic variants with an allele frequency (AF) of less than 0.05%. The findings reveal that a substantial number of these variants, accounting for 31% of the total, are located within the regions spanning from Exons 7 to 11 and exhibit an AF below 0.5%.

**Figure 8:**
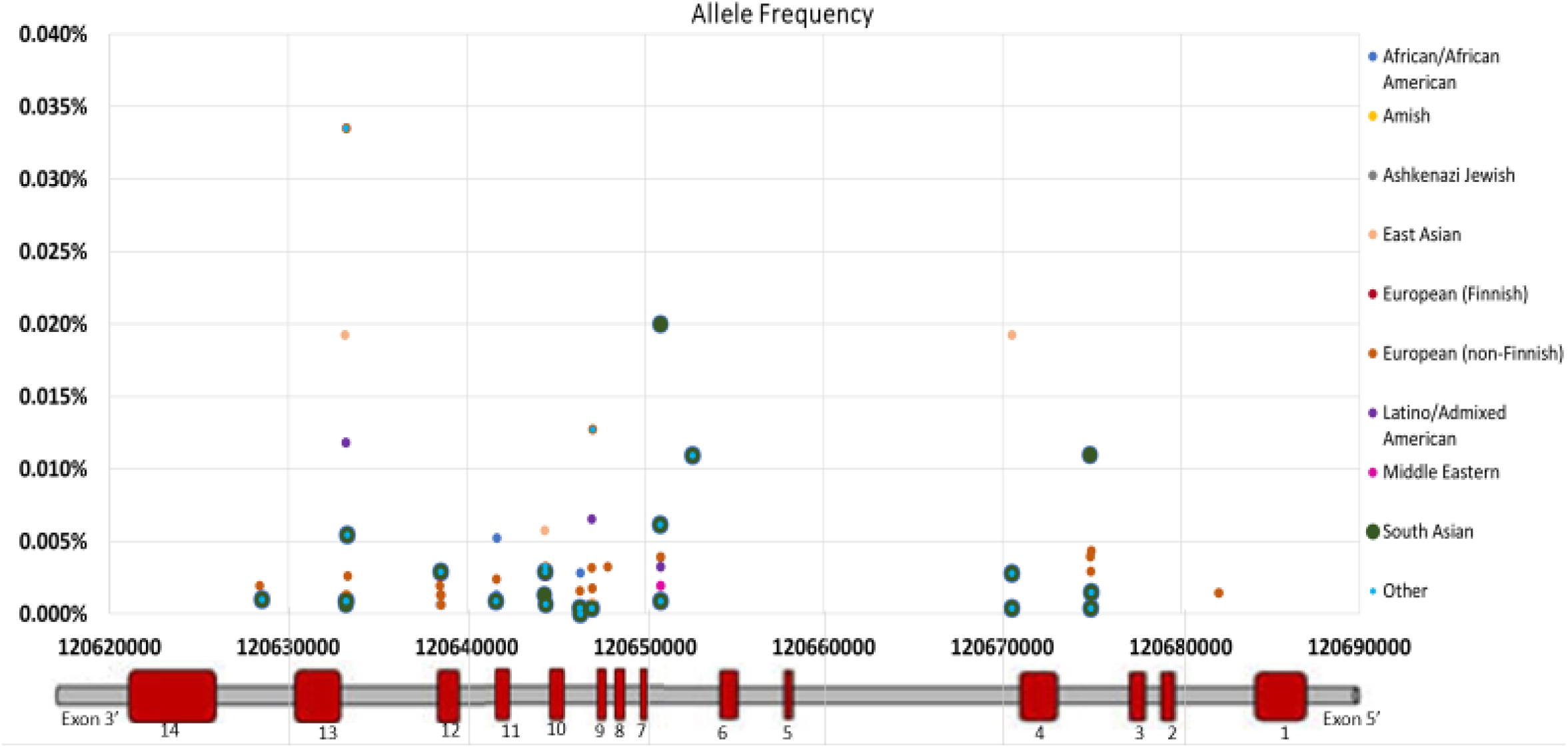
Allele Frequency (AF ≤ 0.0005)-A visual representation of variants with allele frequencies less than 0.0005. 555 HGD variants have allele frequency less than 0.0005. For interpretation purpose, the chromosomal position is referred as Exons here.

A unique observation is that the European (non-Finnish) population stands out as the only ethnic group with variants present in both Exon 1 and Exon 7. Additionally, the African/African American population ranks second in terms of the number of diverse variants present. Distribution of variants in Exon 13 was evenly spread across all ethnic groups. A similar trend can also be seen in Exon 6 and Exon 3.

To investigate allele frequencies in the Indian population, we collected data on 18 variants from Indigene17. Among these variants, c.240A>T, c.372C>T, c.1191A>C, and c.1179T>C were found to be common (Supplement table 4). Our analysis revealed that only a few variants (Supplement table 4) were less frequent in the Indian population, with 45% of these being annotated as benign. Notably, the variant c.1221G>A, which is more common in the global population, was found to be less prevalent in the Indian population.

#### 4.5 Database and features

AKUHub is a dedicated web portal for the HGD gene variants, which has been manually annotated from Literatures and various databases, as per ACMG/AMP guidelines. It contains 1686 exonic variants identified as of December2022. This portal for Alkaptonuria variants has been made publicly available at https://www.zifogenomics.com/AKUHub. The database has the provision for searching in terms of NM/NP ID, chromosome location and pathogenicity of the variants. The filter and search bar aids to retrieve the data from the database with search query (Figure 9 a) and (Figure 9 b).

**Figure 9a):**
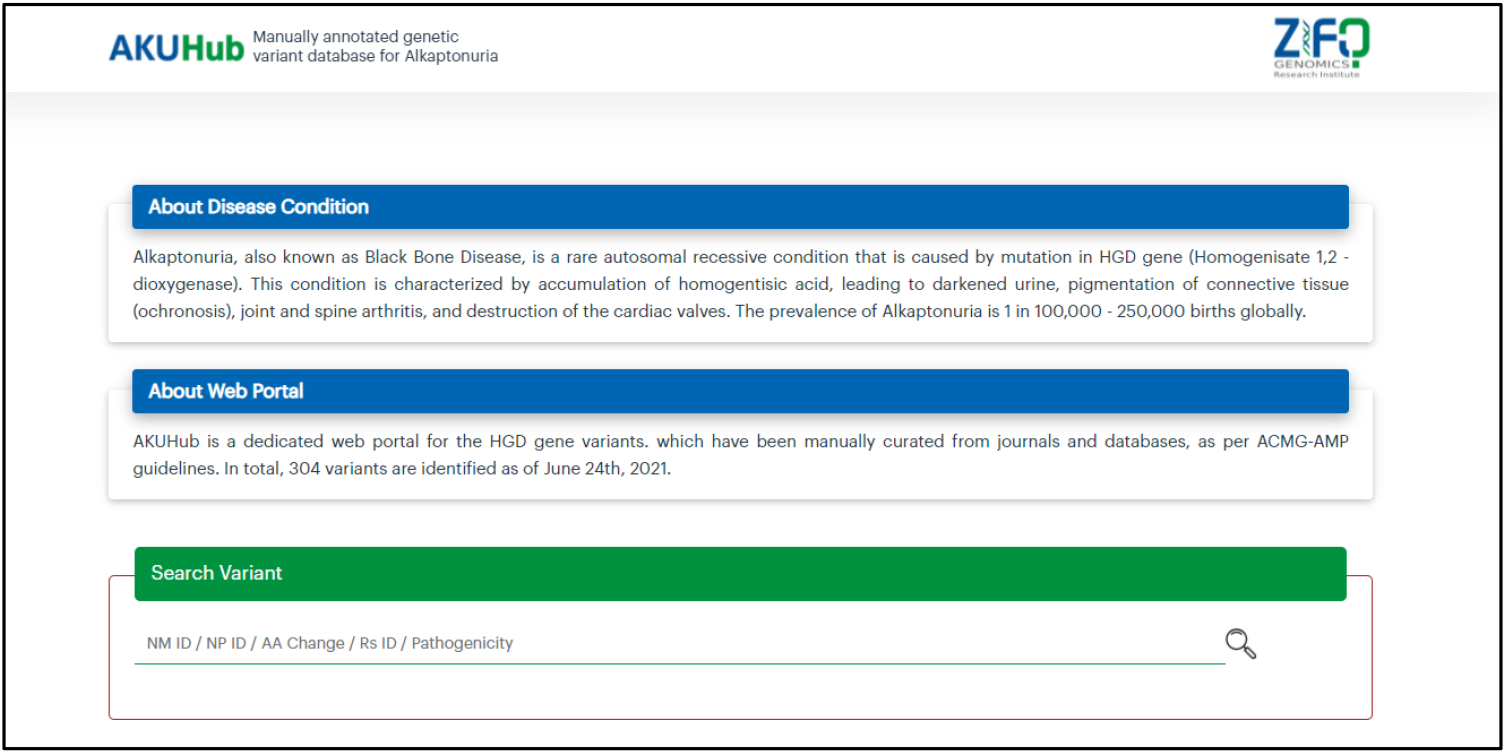
Landing page of the AKU portal- The AKUhub database allows for searching based on HGVS nomenclature, chromosome location, and pathogenicity of the variants.

**Figure 9b):**
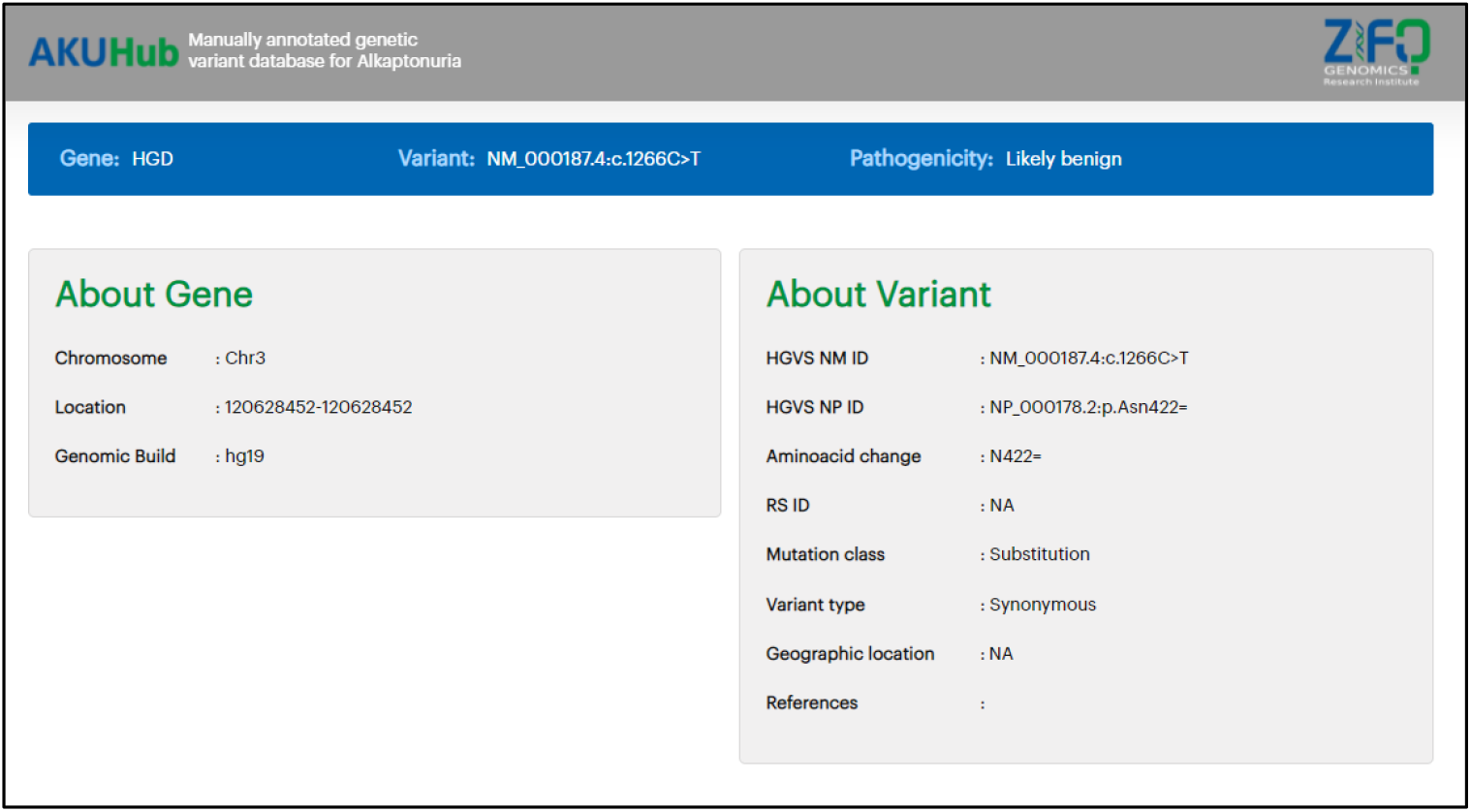
Variant information page of the portal - On the variant information page of the AKU portal information about the variant including its gene name, Chr number, Chr position, HGVS nomenclature, geographic location, references and pathogenicity was displayed.

## 5. Discussion

Variant annotation for rare diseases such as alkaptonuria would assist the ongoing clinical and molecular research. In that context we have manually annotated the exonic variants and created disease specific database called AKUHub for alkaptonuria. It is a publicly available database, which contains all the exonic variants for alkaptonuria in the protein coding region. One of the unique features of this study is that we observed variants (Supplement table 5) which were not previously reported in any of the databases and for which additional evidence may be needed to establish pathogenicity. As of now, these variants are not included in AKUHub. However, they will be added in the future as and when new literatures are published. Currently, these unique variants were considered as variants with uncertain significance. Its details and pathogenicity might be updated in the future based on evolving research and literature evidence. Currently, AKUHub contains 1686 manually annotated variants that would help the genetic counsellors and clinicians to understand the disease pathogenicity. It also helps in finding out the treatment options and counselling the patients with respect to disease severity. AKUHub contains an updated exhaustive list of variants compared to other HGD databases like LOVD HGD Database. As on January 2023 LOVD had around 279 variants reported in various regions of the HGD gene with 72% (∼184) accounting to coding region whereas AKUHub has 1686 exonic variants collected from various data sources including literature which have been manually curated as per the ACMG / AMP guidelines.

During the development of AKUHub, we discovered a few variants that had conflicting results when compared to other well-known sources such as ClinVar (Supplement table 6). We have attempted to resolve inconsistency in annotation by manually reannotating all variants as per the guidelines.

The current focus of AKUHub database is primarily the exonic variants of HGD gene. While on the contrary, for alkaptonuria, there were evidence about the mutations in splice sites and intronic regions. Studies have shown that splice site variants can lead to skipping of exons^31^ and deep intronic mutations result in formation of a cryptic exons^32^. These mutations may have an impact on the HGD gene expression and affect the severity of disease condition. In the line of importance of these regions and association with alkaptonuria, we would like to extend the database with variant information of introns and splice sites.

## Data Availability

All data produced in the present study are available upon reasonable request to the authors

## Acknowledgement

We express our sincere gratitude to Dr. Sridhar Sivasubbu, Senior Principal Scientist, CSIR – IGIB, and Dr. Vinod Scaria, Principal Scientist, CSIR – IGIB, for their valuable guidance throughout the study, which significantly helped in gaining a better understanding of the disease.

Our heartfelt appreciation goes out to our Institute’s Web-Designing and Core annotation team for their contribution in designing AKUHub and interpreting the results, respectively.

We acknowledge Zifo RnD Solutions, Chennai, India, for providing financial support to conduct the study. This research would not have been possible without their assistance.

We extend our gratitude to all the individuals who contributed to this study in various ways, helping us to achieve our objectives.

