## Supplementary Material for "A Compendium of manually annotated genetic variants for Alkaptonuria-AKUHub"

**SUPPLEMENT TABLES OF ALKAPTONURIA**

1. **Supplement Table 1:** List of Software packages and last collected date which is used for the annotation.
2. **Supplement Table 2:** Pathogenic variants with functional analyses done in literature articles.
3. **Supplement Table 3:** Variants with conflicting prediction compared with *in silico* tools.
4. **Supplement Table 4:** Allele Frequency for variants found in India.
5. **Supplement Table 5**: List of variants identified only in literature.
6. **Supplement Table 6**: Pathogenicity comparison of Clinvar and AKUhub.

### Supplement table 1:

| Variants Sources | Last collected date | Total no. of variants |
|---|---|---|
| Leiden Open Variation Database (LOVD) | 12-Dec- 2022 | 256 |
| Indigenomes | 12-Dec- 2022 | 983 |
| Research articles published in English language | 12-Dec- 2022 | 154 |
| ClinVar | 12-Dec- 2022 | 246 |
| gnomAD | 12-Dec- 2022 | 783 |
| dbSNP | 12-Dec- 2022 | 657 |
| GenomAsia | 12-Dec- 2022 | 1405 |
| Mastermind | 12-Dec- 2022 | 1205 |

Supplement table 2:

| NM ID | NP ID | Domain | Functional Assay | PMID |
|---|---|---|---|---|
| NM_000187.4:c.125A>C | NP_000178.2:p.Glu42Ala | HgmA_N | Pathogenic | PMID: 36376482 |
| NM_000187.4:c.178_180delinsGGA | NP_000178.2:p.Trp60Gly | HgmA C | Pathogenic | PMID: 11001939 |
| NM_000187.4:c.185A>G | NP_000178.2:p.Tyr62Cys | HgmA_N | Pathogenic | PMID: 36376482 |
| NM_000187.4:c.289_291delinsGGA | NP_000178.2:p.Trp97Gly | HgmA C | Pathogenic | PMID: 11001939 |
| NM_000187.4:c.365_366delinsTA | NP_000178.2:p.Ala122Val | HgmA_N | Pathogenic | PMID: 30737480 |
| NM_000187.4:c.481G>A | NP_000178.2:p.Gly161Arg | HgmA C | Pathogenic | PMID: 30737480 |
| NM_000187.4:c.566G>T | NP_000178.2:p.Ser189Ile | HgmA C | Pathogenic | PMID: 11001939 |
| NM_000187.4:c.688C>T | NP_000178.2:p.Pro230Ser | HgmA_N | Pathogenic | PMID: 36376482 |
| NM_000187.4:c.1081G>A | NP_000178.2:p.Gly361Arg | | Pathogenic | PMID: 36376482 |
| NM_000187.4:c.1102A>G | NP_000178.2:p.Met368Val | HgmA C | Pathogenic | PMID: 11001939 |
| NM_000187.4:c.1112A>G | NP_000178.2:p.His371Arg | HgmA C | Pathogenic | PMID: 11001939 |

Supplement Table 3:

| NMID | NPID | In silico Prediction | ACMG curated |
|---|---|---|---|
| NM_000187.4:c.832_834delinsGAT | NP_000178.2:p.Asn278Asp | Benign | Likely Pathogenic |
| NM_000187.4:c.920G>A | NP_000178.2:p.Arg307His | Benign | Likely Pathogenic |
| NM_000187.4:c.923C>G | NP_000178.2:p.Pro308Arg | Benign | Likely Pathogenic |
| NM_000187.4:c.976G>A | NP_000178.2:p.Asp326Asn | Benign | Likely Pathogenic |

Supplement Table 4:

| NMID | Allele frequency (India) | Allele frequency (Global) |
|---|---|---|
| NM_000187.4:c.240A>T | >1% | >5% |
| NM_000187.4:c.372C>T | >1% | 5.300% |
| NM_000187.4:c.1191A>C | >1% | 2.100% |
| NM_000187.4:c.1179T>C | >1% | 1.11% |
| NM_000187.4:c.920G>A | 0.05% | 0.003% |
| NM_000187.4:c.752G>A | 0.05% | 0.001% |
| NM_000187.4:c.685A>G | 0.05% | 0.011% |
| NM_000187.4:c.501C>T | 0.05% | 0.009% |
| NM_000187.4:c.158G>A | 0.05% | 0.004% |
| NM_000187.4:c.111C>T | 0.05% | 0.010% |
| NM_000187.4:c.1221G>A | 0.05% | 0.150% |
| NM_000187.4:c.237C>T | 0.05% | 0.042% |
| NM_000187.4:c.19A>C | 0.05% | 0.001% |
| NM_000187.4:c.365C>T | 0.10% | 0.020% |
| NM_000187.4:c.307C>A | 0.20% | 0.016% |
| NM_000187.4:c.175del | 0.15% | 0.011% |
| NM_000187.4:c.141G>A | 0.10% | 0.012% |

Supplement Table 5:

| Literature Variants | Pathogenicity | Exon Number | Article Referred In (PMID) |
|---|---|---|---|
| NM_000187.4:c.342delA | VUS | Exon 3 | PMID: 18945288 |
| NM_000187.4:c.648G>A | VUS | Exon 8 | PMID: 12051967 |
| NM_000187.4:c.855C>T | VUS | Exon 11 | PMID: 12051967 |
| NM_000187.4:c.855C>A | VUS | Exon 11 | PMID: 16085442 |
| NM_000187.4:c.909A>G | VUS | Exon 10 | PMID: 16085442 |
| NM_000187.4:c.1269A>G | VUS | Exon 13 | PMID: 16085442 |
| NM_000187.4:c.1279A>G | VUS | Exon 13 | PMID: 16085442 |
| NM_000187.4:c.819delG | VUS | Exon 10 | PMID: 16085442 |
| NM_000187.4:c.1183delT | VUS | Exon 13 | PMID: 16085442 |
| NM_000187.4:c.1388G>A | VUS | Exon 14 | PMID: 16085442 |
| NM_000187.4:c.178T>A | VUS | Exon 4 | PMID: 16085442 |
| NM_000187.4:c.594A>T | VUS | Exon 6 | PMID: 16085442 |

Supplement Table 6:

| HGVSc | HGVSp | Variant Type | AKUHub's Clinical Significance | Clinvar's clinical significance |
|---|---|---|---|---|
| NM_000187.4:c.1027A>C | NP_000178.2p.Met343Leu | Missense | VUS | Likely benign |
| NM_000187.4:c.1081G>A | NP_000178.2p.Gly361Arg | Missense | Pathogenic | Uncertain significance |
| NM_000187.4:c.1266C>T | NP_000178.2p.Asn422= | Synonymous | Likely Benign | Uncertain significance |
| NM_000187.4:c.157C>T | NP_000178.2p.Arg53Trp | Missense | Likely Pathogenic | Uncertain significance |
| NM_000187.4:c.221A>T | NP_000178.2p.Glu74Val | Missense | VUS | Benign/Likely benign |
| NM_000187.4:c.260A>C | NP_000178.2p.Glu87Ala | Missense | VUS | Benign |
| NM_000187.4:c.307C>A | NP_000178.2p.Pro103Thr | Missense | VUS | Conflicting - VUS / Likely Benign |
| NM_000187.4:c.413G>A | NP_000178.2p.Cys138Tyr | Missense | VUS | Likely pathogenic |
| NM_000187.4:c.48A>G | NP_000178.2p.Ser16= | Synonymous | Likely Benign | Uncertain significance |
| NM_000187.4:c.567C>G | NP_000178.2p.Ser189Arg | Missense | Likely Pathogenic | Uncertain significance |
| NM_000187.4:c.709C>T | NP_000178.2p.Arg237Cys | Missense | Likely Pathogenic | Uncertain significance |
| NM_000187.4:c.710G>A | NP_000178.2p.Arg237His | Missense | Likely Pathogenic | Uncertain significance |
| NM_000187.4:c.752G>A | NP_000178.2p.Gly251Asp | Missense | Likely Pathogenic | Uncertain significance |
| NM_000187.4:c.753C>T | NP_000178.2p.Gly251= | Synonymous | VUS | Likely pathogenic |
| NM_000187.4:c.919C>T | NP_000178.2p.Arg307Cys | Missense | VUS | Likely benign |
| NM_000187.4:c.920G>A | NP_000178.2p.Arg307His | Missense | Likely Pathogenic | Uncertain significance |
| NM_000187.4:c.923C>G | NP_000178.2p.Pro308Arg | Missense | Likely Pathogenic | Uncertain significance |
